# Transferability of Psychological Interventions from Disaster-Exposed Employees to Healthcare Workers Working during the COVID-19 Pandemic

**DOI:** 10.1101/2021.10.28.21265604

**Authors:** Sean Treacy, Shane O’Donnell, Blánaid Gavin, Tamara Schloemer, Etain Quigley, Dimitrios Adamis, Fiona Mc Nicholas, John C. Hayden

**Author notes:** Correspond Author, Corresponding Author, Corresponding Author Address: School of Pharmacy and Biomolecular Sciences, RCSI University of Medicine and Health Sciences, 111 St Stephen’s Green, Dublin 2, Ireland. No funding was received for this study. The study does not require ethical approval as no human or animal data is used.

## Abstract

**Background:** The COVID-19 Pandemic had a significant negative impact on the mental health of healthcare workers. Evidence-based interventions that could be used to mitigate this impact are lacking in the literature. This review aims to evaluate psychological interventions used for employees following previous disasters and assess the transferability of these interventions to a healthcare setting during the COVID-19 pandemic.

**Methods:** Intervention information from a previously published systematic review of the literature published up to 2015 was extracted, and an additional search of studies published from 2015-2020 was conducted. Studies were assessed for transferability using a checklist derived from the PIET-T process model.

**Results:** Interventions from eighteen studies were assessed for transferability (including three studies identified in an updated literature search). Interventions established as most transferable included resilience training, meditation/mindfulness interventions, and cognitive behavioural therapy. Psychological debriefing was transferable but as it is contrary to current recommendations is not deemed appropriate for adoption.

**Implications:** Several existing interventions have the potential to be utilised within the COVID-19 context/pandemic. More research needs to be undertaken in this area to assess these interventions upon transfer.

## Introduction

Much of the initial media coverage about the COVID-19 pandemic focused almost solely on the physical consequences of the virus, and yet only in the latter stages has the considerable psychological fallout been highlighted. Those particularly at risk, both psychically and psychologically, are healthcare workers (HCWs) working on the front line. HCWs working in direct contact with affected patients during previous virus outbreaks have been shown to be at increased risk of post-traumatic stress and psychological distress.^1^ Similar findings have been seen with the COVID-19 pandemic, with increased levels of anxiety, burnout, depression and sleeping problems reported among HCWs.^2,3^ Risk factors associated with psychological distress and negative mental health outcomes included a lack of appropriate Personal Protective Equipment (PPE) and unclear communication from hospital management.^1^

Possible mental health interventions such as psychological debriefing and psychological first aid have been suggested as potentially useful in managing the psychological impact of the pandemic^4^; but, to our knowledge, there have been no reviews exploring the repurposing and transferability of interventions used during previous disasters in relation to their applicability to the current pandemic. Outside of psychological interventions developed for HCWs, for example, resilience training used during previous pandemics, the broader scope of all employee-based psychological interventions also merits exploration, for example, supports following natural disasters.

This study designed and utilised a transferability checklist using Schleomar and Schröder-Bäck’s (2018) Population, Intervention, Environment, Transfer and Transferability process model (PIET-T).^5^ The PIET-T process model is a model designed to assess whether health interventions can be transferred from the “primary context” (i.e. the context of the intervention as it was performed in the original study) to the “target context” (i.e. the context that the intervention is aimed at being performed in). In this study, the target context was ‘Frontline healthcare workers in a hospital setting in Ireland’. Ireland was chosen specifically as it is the context with which the analysts are most familiar. The primary and target contexts are evaluated in terms of their population (P) and environment (E) and the intervention is then assessed for whether it can be transferred from one context to the other.

The aim of this article is to evaluate the transferability of psychological interventions that have been previously used with employees after a disaster situation to a hospital setting during the COVID-19 pandemic.

## Methods

### Identifying previous literature

A secondary analysis of the studies identified in a 2018 systematic review^6^ by Brooks et al (2018) of interventions for the psychological impacts on disaster-exposed employees was performed to find suitable employee psychological interventions. The 2018 review search terms were also repeated and adapted from the period of 01/01/2015 to 26/06/2020. The search strategy adaptation included the addition of two new search terms in the “disaster” section: “COVID-19” and “coronavirus” EMTREE thesaurus terms. The full search strategy is reported in Supplementary Table 1. Search results were collected on Endnote X9 and duplicates were removed. Studies were initially screened by title then abstract and then by full text by one author (ST).

The inclusion criteria were similar to those used in a previous review (Brooks et al., 2018). These criteria excluded studies which were not peer reviewed and/or in the English language. Further inclusion criteria encompassed a requirement for the study to have included employed participants (defined as any occupational group; any group of people that work together within hierarchical systems to achieve some sense of group aim). In addition included studies needed to involve a disaster, the definition of which relied on the authors own characterisation of a disaster. As such, a variety of natural and ‘manmade’ events were included such as earthquakes, a hurricane, combat exposure, a robbery, an explosion, and a train crash. A further criterion for inclusion was that the study incorporated a psychological intervention that aimed to help employees cope during or after a stated disaster. Studies were excluded if they did not evaluate the effects of a psychological intervention. Extracted data was tabulated under the headings: Study, Journal, Country, Disaster, Design, Participants and role of participants, Intervention, Outcomes Assessed, Results. To ensure consistency with the previous review, the same quality appraisal tool was used. The tool focused on three areas for evaluation: Study design, data collection and methodology and analysis and interpretation of results.

### Assessment of transferability

The authors created a checklist using the PIET-T process model as its basis with questions grouped into assessments of Population (P), Intervention (I), Environment (E), Transfer (T). All studies were initially screened to establish whether transfer was possible with the question “Is it plausible to transfer this intervention to the target context?”. Studies were analysed using a checklist containing nine questions, with assessment responses as follows: (i) yes (Y), (ii) no (N), (iii) yes with adaptation (A) (iv) unknown (U). Two independent scores were conducted (ST and JH). Where there was a discrepancy of more than two Yes responses, a third independent scorer (SO’D) assessed the intervention, with average of the three scores rounded to the nearest integer being used in these cases. This resulted in a transferability rating on the basis number of yes responses assigned per study. It is important to note that by using this model a study may be deemed to have a high transferability potential to frontline HCWs during the pandemic, but this does not characterise the potential effectiveness of the intervention were it successfully transferred.

## Results

The updated search yielded 7,433 records, three studies of which were eligible for inclusion in this review. The flow diagram for the search is shown in Fig. 1. Two of the studies were longitudinal and one of the studies was mixed methods with both a quantitative and qualitative assessment of the intervention. Unlike the previous review^6^ no studies evaluated any pre-disaster interventions with employees who had not yet experienced a disaster. All three additional interventions involved post-disaster interventions. Extracted data from each study is shown in Table 1.

**Fig. 1.**
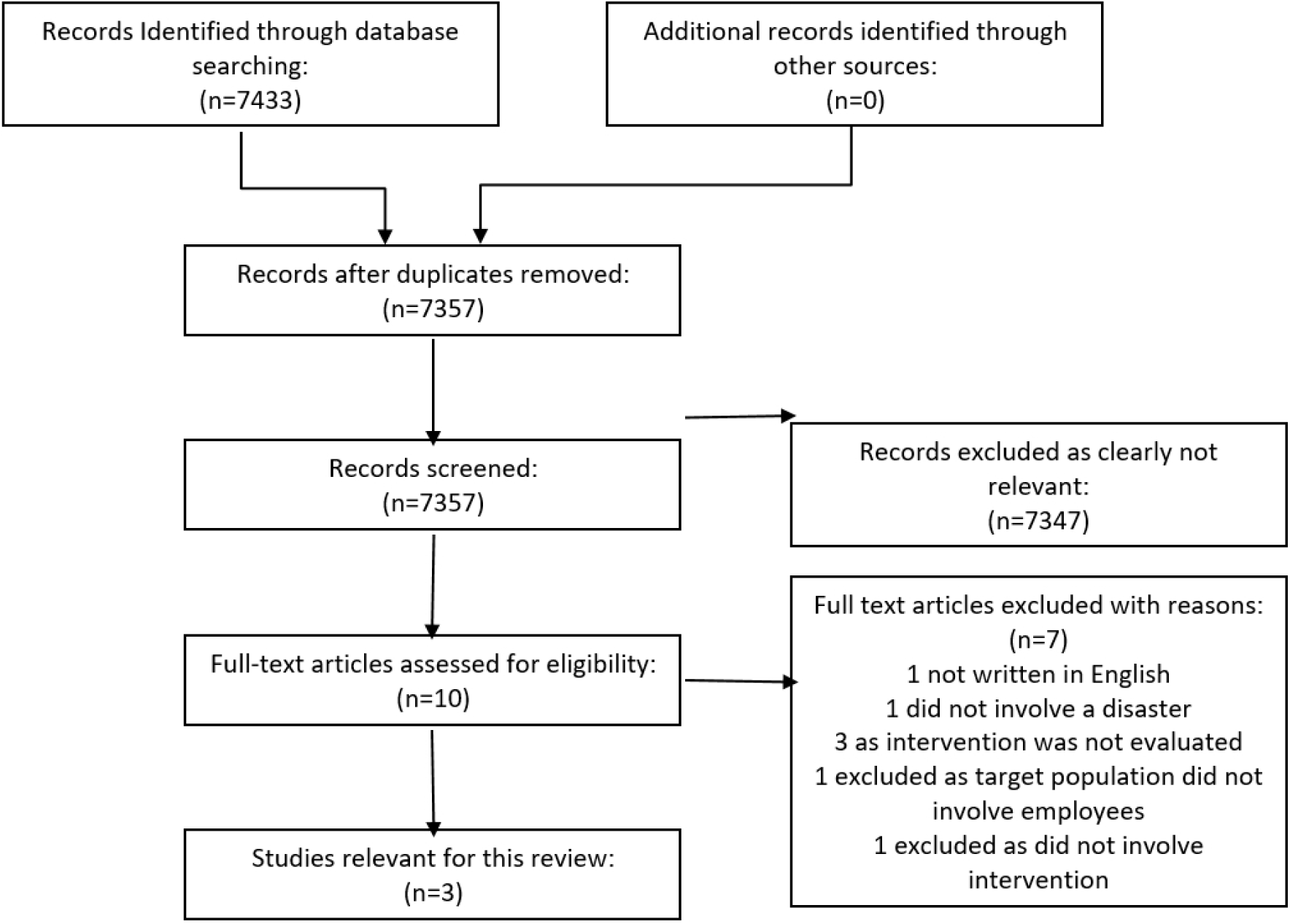
Prisma Flow Diagram of Literature Search

**Table 1.** Characteristics of new studies included in the review

| Study | Country | Disaster | Design | Participants | Intervention | Outcomes Assessed | Quality assessment | Results |
| --- | --- | --- | --- | --- | --- | --- | --- | --- |
| Iwakuma et al. (2017)<br>Effects of Breathing-Based Meditation on Earthquake-Affected Health Professionals<sup>7</sup> | Japan | Great East Japan Earthquake (2013) | Mixed-methods experimental study – Survey completed pre and post meditation session | Health professionals (n=17) | A 45-minute breathing-based meditation session | Temporary moods of depression, anger, fatigue, vigour, strain, and confusion. A qualitative component included open ended questions | 73.3% | All scales showed statistically significant improvements<br>anger: $z(17) = -2.38$, $P = .018$;<br>confusion: $z(17) = -3.3$, $P = .001$;<br>depression: $z(17) = -2.83$, $P = .005$;<br>fatigue: $z(17) = -3.3$, $P = .001$;<br>strain: $z(17) = -2.94$, $P = .003$;<br>vigour: $z(17) = -2.59$, $P = .01$ |
| Jones et al. (2017)<br>Trauma Risk Management (TRiM): Promoting Help Seeking for Mental Health Problems Among Combat-Exposed U.K. Military Personnel<sup>8</sup> | United Kingdom | Combat Exposure | Longitudinal study - Records of TRiM activity during a U.K. military deployment in Afghanistan were linked to contemporaneous survey data assessing | U.K. military personnel (n= 638) | Trauma Risk Management (TRiM) - a peer-led, occupational mental health support process that aims to identify and assist U.K. military personnel with persistent | Mental health and help-seeking outcomes were compared between a nonexposed, non-TRiM sample (n = 161), an exposed, non-TRiM sample (n = 149), and an exposed, | 100% | TRiM recipients had significantly greater adjusted odds of seeking help from formal mental health services than exposed non-TRiM study participants. At both the baseline and follow-up points, TRiM recipients' |

Transferability psychological interventions In Covid 19
|  |  |  | mental health and combat experiences |  | mental ill health related to potentially traumatic events (PTEs) | TRiM-recipient sample (n = 328) |  | functional impairment levels were not significantly different to exposed non-TRiM participants. |
| --- | --- | --- | --- | --- | --- | --- | --- | --- |
| Ke et al. (2017) Posttraumatic Psychiatric Disorders and Resilience in Healthcare Providers following a Disastrous Earthquake: An Interventional Study in Taiwan<sup>9</sup> | Taiwan | Earthquake | Longitudinal study – baseline data collected on site after disaster and 1 month after disaster follow up | HCWs (n=67) including doctors (n=32) and nurses (n=35) | On site debriefing courses and mini lectures for the Health care providers | Recurrent and intrusive distressing recollections of the event, tachycardia; muscle tension; difficulty relaxing; difficulty falling or staying asleep; feeling fear; feeling guilty; needing help after the medical response; and needing to talk with someone in private. | 81.3% | The incidence of post-traumatic psychiatric disorders was 16.4% (11/67) in all the HCPs. After the intervention, the follow up questionnaire 1 month later revealed no symptoms among the total HCPs |

### New studies since 2015

A summary of the three additional studies identified is reported in Table 1. Overall quality of studies was relatively high with no study scoring below 70% based on the quality appraisal tool designed by the authors of the previous review.^6^

The first study^7^, by Iwakuma et al. 2017, assessed the effectiveness of a 45-minute breathing-based meditation on HCWs following an earthquake. Participants were assessed using a temporary mood scale which measured levels of depression, anger, fatigue, vigour, strain, and confusion. A qualitative evaluation was also conducted where participants were asked to give an account of their subjective experience of the meditation (Iwakuma et al., 2017). All temporary mood scales showed statistically significant improvement after the meditation session (anger: z (17) = -2.38, P = .018; confusion: z (17) = -3.3, P = .001; depression: z (17) = -2.83, P = .005; fatigue: z(17) = -3.3, P = .001; strain: z (17) = -2.94, P = .003; vigour: z (17) = -2.59, P = .01). Steps Coding and Theorization (SCAT) was used to assess the qualitative component of the study, and participants reported sensations such as “emancipation from chronic and bodily senses”; “holistic sense”: “transcending mind-body”.

The second additional study^8^ identified, Jones et al. 2017, was a United Kingdom based study assessing the intervention Trauma Risk Management (TRiM) - a peer-led, occupational mental health support process that aims to identify and assist U.K. military personnel with persistent mental ill health related to potentially traumatic events (PTEs). Mental health and help-seeking outcomes were compared between a non-exposed group, an exposed group and an exposed group not receiving the TRiM intervention. Following the intervention, TRiM recipients had significantly greater adjusted odds of seeking help from formal mental health services than exposed non-TRiM study participants. At both the baseline and follow-up points, TRiM recipients’ functional impairment levels were not significantly different to exposed non-TRiM participants. Stigma and perceived barriers to care levels were not significantly different between the exposed non-TRiM and TRiM recipient groups.

The third study^9^, Ke et al. 2017, assessed the impact of on-site debriefing and mini lectures for health care providers following an earthquake. Participants were assessed for post-traumatic stress symptoms (such as recurrent and intrusive distressing recollections of the event, tachycardia; muscle tension; difficulty relaxing etc.) following the event using a questionnaire. This same questionnaire was given one month later as a follow up. Every participant received the psychological debriefing and mini lectures from trained psychologists and psychiatrists. The study on psychological debriefing courses and mini lectures following an earthquake revealed that the incidence of post-traumatic psychiatric disorders was 16.4% (11/67) in all the health care professionals (HCPs). After the intervention, the follow up questionnaire one month later revealed no symptoms among the total HCPs. This study did not have a comparator group of participants unexposed to the intervention.

### Assessment of Transferability

Of the 15 studies reported in a previous review^6^ and the three studies^7-9^ identified during this updated literature search, 17 studies^7-24^ were assessed for transferability. A study^25^ by Eid et al. (2004) was excluded from assessment of transferability based on the plausibility-screening question as it took place on a submarine and the intervention involved the simulation of a submarine manoeuvre. Boscarino et al 2005; Boscarino et al. 2006 were also assessed together as both studies evaluated the same intervention (brief on-site mental health interventions) occurring at the same primary context (World trade centre disaster).^20,21^ Transferability ratings for the remaining 16 studies are reported in Table 2. The mean transferability score was 4.875 out of a possible nine. Interventions that involved HCWs, mental health workers or emergency responders tended to score highly due to the similarities of these populations with the target population. Interventions that took place in a hospital setting tended to score highly. Questions such as “Is the evidence base of the intervention appropriate for the target context” and “Is the Intervention content appropriate for the target context” received a “No” response in the majority of cases. Many studies did not contain controls or did not involve adequate follow up. In many cases the intervention itself was poorly described and therefore difficult to replicate in a different context. Wu et al. 2012 was the only study to refer to how the intervention might be transferred.^18^ In this study, the intervention is described as being “developed according to the actual characteristics of Chinese military workers” and cautioned that there may be “too many differences found between military and non-military organisations.

**Table 2:**
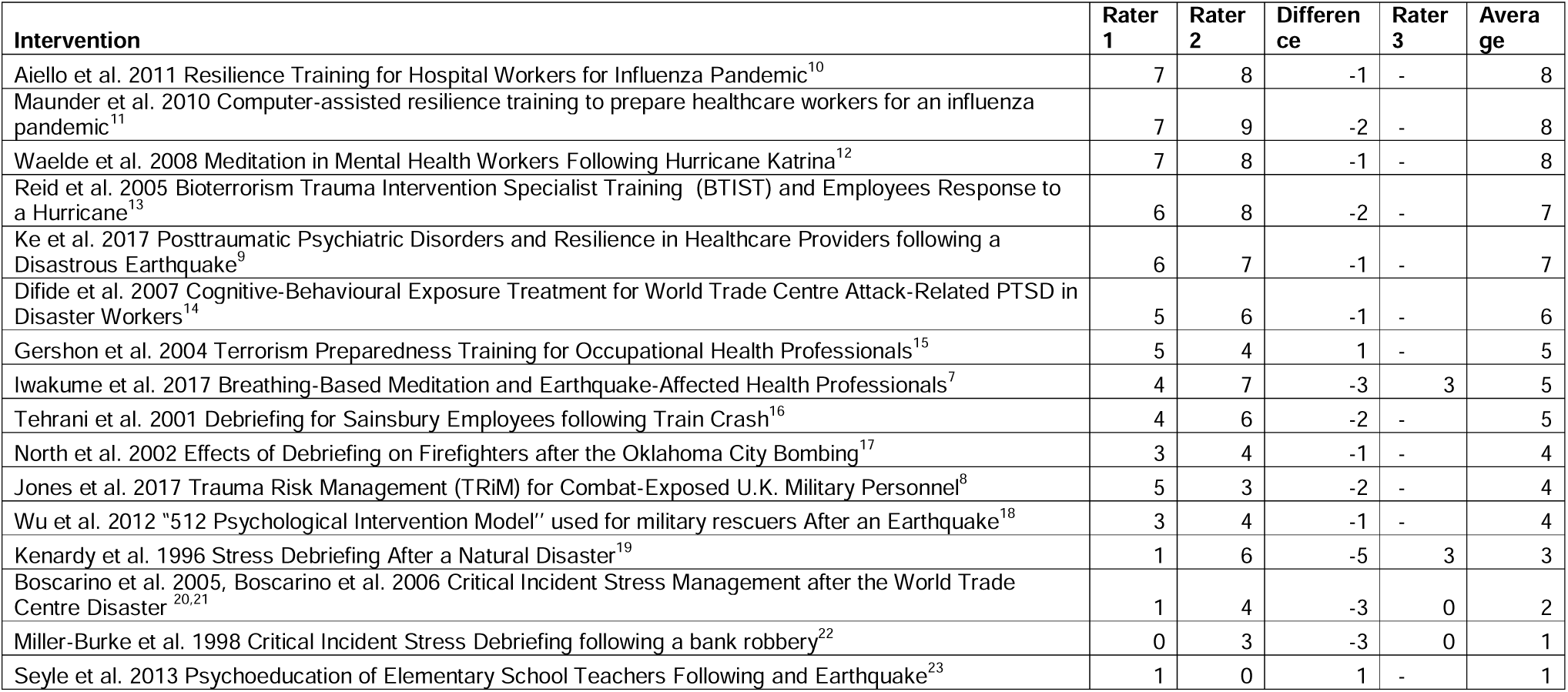
Transferability of interventions rating scores

### Interventions with the highest transferability score

The five studies that received the highest score based on the transferability checklist included three studies^10,11,13^ that involved mental health training courses for HCWs, one study^12^ that evaluated the impact of meditation on HCWs following a hurricane and one study^9^ that evaluated the impact of on-site debriefing courses following an earthquake.

Maunder et al. 2010 assessed the efficacy of a computer aided resilience-training course for HCWs in Mt. Sinai Hospital Toronto, Ontario, Canada in preparation for the H1N1 influenza pandemic of 2009.^11^ Different course lengths were randomised between participants to evaluate what the optimum length of time a resilience-training course should take (1.5hrs, 3hrs, 4hrs). One hundred and fifty eight HCWs were randomly assigned to courses of different lengths. Participants received paid educational time for participating. The primary and target contexts were very similar with this study as it took place during a previous pandemic in a hospital setting. The study described the characteristics of the training course and could potentially be used as a guide for the development of a novel training course that is more relevant to the current pandemic. Key results of this study include the fact that partaking in the course increased the degree of confidence participants felt towards working during the pandemic, increased the confidence participants had towards in training and support the participants received and decreased interpersonal problems (socially inhibited, non-assertive, overly accommodating etc.) It also showed that a three-hour course length seemed to be the most efficacious and that longer course times lead to increased rates of dropout. The study did not directly measure improvements in pandemic related stress because of the intervention and there was no follow up during an influenza outbreak so efficacy in that context could not be measured.

Aiello et al. (2011) also studied the impact of a resilience training session in preparation for the 2009 H1N1 influenza pandemic in Mt. Sinai Hospital Toronto, Ontario Canada.^10^ One thousand two hundred and fifty HCWs received training with 1020 (82%) participants returning evaluations. The intervention involved training in coping mechanisms using the “Folkman and Greer’s model” of coping and encouraged group discussion and participation. The intervention was deemed to be highly transferable as it took place in a hospital during a previous pandemic. There was limited detail regarding the intervention itself. There was no control group and no follow up examination during an influenza outbreak. Psychological parameters were not measured, and efficacy was evaluated based on participant feedback, with 76% of participants reported being better able to cope after the training sessions. The study itself cautions against generalising its results as a large proportion of hospital staff worked during a previous SARS outbreak in the hospital and that this may have made the intervention of a higher perceived importance in the staff relative to other hospitals.

Reid et al. (2005) studied the efficacy of Bioterrorism Trauma Intervention Specialist Training (BTIST) curriculum on public health workers following four hurricanes that struck Florida during a seven week period.^13^ A web-based survey was performed on BTIST participants that evaluated how the training enhanced their ability to understand traumatic stress reactions and perform psychological defusings and debriefings. This intervention mainly focused on training workers to apply disaster mental health interventions to survivors of disasters, but one aspect of the training involved “compassion fatigue resiliency training”, the components of which were not described in the study. The evidence-base for this intervention is limited. The study involved sending a survey over email with a 40% response rate and 53 participants. A thorough description of the intervention itself and the components of the compassion fatigue resilience training are not included in the published manuscript. The efficacy of compassion fatigue resilience training was also not measured.

Waelde et al. (2008) evaluated the effects of a four-hour mindfulness workshop followed by eight weeks of mindfulness practice at home on mental health workers following Hurricane Katrina in New Orleans.^12^ During the workshop, participants completed a self-reported baseline assessment and mail in assessments three and eight weeks postworkshop. Twenty mental health workers participated, five of whom did not complete the postworkshop assessments. Ninety four percent of participants reported feeling “some-what better” or “much better” than before the study after the intervention. The slopes of change for total PTSD and state anxiety symptoms were correlated with the total number of minutes meditated across the eight weeks (r = -40, p < 0.05, and r = -38, p < 0.05 respectively). An increased number of minutes spent meditating was associated with greater improvements on PTSD and anxiety symptoms. The study lacked a control group and only 15 participants completed the post baseline assessments.

Ke et al. 2017 evaluated the effects on-site debriefing courses, mini lectures, muscle relaxation techniques on health care providers following an earthquake in Taiwan.^9^ Participants completed a questionnaire that assessed symptoms of post-traumatic stress disorder immediately after the onsite intervention and at one month follow up. After the initial intervention, 16.4% of participants had at least one symptom, while no participant had at least one symptom one month after the intervention. There was no control group for this study and the sample size constituted 67 participants.

## Discussion

Psychological interventions that hospitals could readily employ to improve the mental health of their frontline HCWs during a pandemic are largely lacking in the literature. This study identified psychological interventions previously implemented following other disasters and assess whether these interventions could be transferable to frontline HCWs in a hospital setting in Ireland during the COVID-19 outbreak. The most transferable Interventions were based upon resilience training, psychological debriefing, meditation/mindfulness or multidimensional. While evidence for effectiveness of these interventions is limited, adoption of these interventions may be deemed appropriate under the discretion of healthcare management. Further adaptations may enable delivery remotely to facilitate physical distancing rules and enable widespread economical delivery.

Three studies assessed the effectiveness of resilience training prior to an infectious disease outbreak in a hospital setting, each receiving high scores in transferability.^10,11,13^ Resilience has been defined as the ability to adapt and effectively cope with adversity, life stressors and traumatic events.^25^ The evidence for resilience training sessions has been evaluated in a Cochrane review which found that there was evidence that HCWs receiving resilience training reported higher levels of resilience, lower levels of depression and stress compared to controls.^26^ Learned resilience has emerged as a psychological intervention strategy to prepare HCW for occupation-related stressors. The individual is taught how to pre-empt likely stressors, possible reactions, and symptoms, and developing behavioural and cognitive coping strategies. Enhanced self-care practices are also described as fundamental to developing resilience.^27^ Maunder et al. (2010) and Aiello et al (2011) evaluated resilience training which utilised relaxation techniques and helped participants identify more effective coping mechanisms^10,11^, while the intervention described by Reid et al. (2005), participants received compassion fatigue resiliency training^13^. These interventions were originally studied in a hospital setting in the original study context, suggesting they are more readily transferable than non-hospital-based interventions. In particular, Maunder’s intervention already exists as a computer delivered course with participants who under-utilized coping via problem-solving or seeking support or over-utilized escape-avoidance experiencing improved coping.^11^ A course of seven sessions (158 minutes) was associated with positive outcomes in a randomised study design with acceptable drop-out rates. The transferable studies of Reid et al. (2005) [resilience and compassion fatigue training] and Aeillo et al. (2011) [resilience training] were both delivered in person, but remote delivery could be explored.^10,13^ Multimedia versions of components of the intervention already exist for Reid’s intervention, and an outline curriculum is available for adaptation.^13^

Two studies evaluate a mindfulness-based approach to mitigating psychological distress to disaster exposed HCWs.^7,12^ Waelde’s intervention was rated highly for transferability.^12^ The primary context and the target context are very similar for both of these studies as they both involve HCWs. Waelde studied psychological outcomes of an eight week mindfulness course and found a negative association between the rates of post-traumatic stress disorder (PTSD) and anxiety and the number of minutes meditated. While there are many forms of meditation and mindfulness, those with an evidence base such as mindfulness-based stress reduction (MBSR) are of particular interest as to potential transferability within the context described. Mindfulness have shown improvements in measures of anxiety, depression, and pain scores. Structural and functional brain changes have been demonstrated in the brains of people with a long-term traditional meditation practice, and in people who have completed a MBSR programme.^28^ While more evidence is necessary to evaluate the impact this intervention could have during the COVID-19 pandemic, the low cost and minimum workload required for implementation could enable more hospitals to adopt this intervention. Mindfulness apps could be provided to HCWs by hospital management and participants could practice in accordance with their own schedule.

Difede et al. (2007) evaluated the potential use of cognitive behavioural therapy in disaster workers following the World Trade Centre attacks.^14^ It is the sole randomised control trial of all the studies evaluated in this review. The intervention group was shown to have lower Clinician Assessed PTSD Scores (CAPS) than the treatment as usual group following 12 weekly sessions of cognitive behavioural therapy. Use of online video calls as a substitute for in person therapy would allow this intervention to be compatible with social distancing. However, under a “screen and treat” approach, ideally, participants at risk of developing PTSD will need to be identified first and then treatment would adhere national or local treatment guidelines.

Seven studies employed forms of psychological debriefing as a way to decrease the probability of developing post-traumatic stress symptoms following a potentially traumatic event.^16-22^ These interventions tended to take place in primary contexts that were very different to this study’s target context and thus tended to score below average for transferability in this study. Psychological debriefing is a treatment that has come under criticism. A systematic review published in 2002 reports that psychological debriefing has the potential to have negative effects and instead recommends a “screen and treat” model as an organisation level psychological intervention.^29^ Guidelines published in 2018 by the National Institute for Health Care Excellence (NICE) recommend against the use of psychological debriefing (2018).^30^ The review that this current study is based on also advises against the implementation of psychological debriefing due to the potential negative effects.^6^ While psychological debriefing could be implemented into a healthcare setting for HCWs exposed to a potentially traumatic event during the pandemic, it is contrary to current policy recommendations.

This review aimed to identify interventions used during previous disasters to allow healthcare managers consider repurposing existing interventions for rapid deployment in the current pandemic. However, strengths and limitations of our approach need to be considered. A checklist was used in this derived from the “PIET-T Process model” which was designed specifically to examine the scope for the potential transferability of healthcare interventions from one context to another. While it is a strength to have a theoretically informed approach, there are no established cut-offs for transferability. We chose to focus on those interventions that were amongst the most highly rated and tried to minimise subjectivity by using multiple scorers. The limited research in the area meant we had to broaden our scope to all workers and not just healthcare workers. Extracting intervention information from a previous systematic review^6^ allowed for a more thorough search of the literature to be conducted. Studies not written in English were excluded from the search and this may have led to important studies being left out of the review. Only one database was searched for updated relevant literature from 2015 onwards. Only one study gave any information about how the intervention could be transferred to a different context. The target context of this study included frontline HCWs in a hospital setting in Ireland due to the analysts involved being more familiar with this healthcare system, this may limit the generalisability of our findings. Interventions were generally poorly described and therefore replication of the original intervention may be difficult. A thorough examination of robustness of evidence as to the effectiveness of each intervention was beyond the scope of this review and needs to be considered before intervention adoption. Our search was up to June 2020, so the latest research is not included. Clearly, this area continues to evolve rapidly; however, the focus of our review was on the possibility of repurposing pre-pandemic interventions.

## Conclusions

There remains a lack of literature regarding evidence based optimum psychological interventions for employees following a disaster. The results of the PIET-T process model-based checklist designed for this review could be used by decision makers to assess whether specific interventions could be transferred to their target setting and used during COVID-19. A number of interventions are worthy of consideration for adoption. Computer-assisted resilience training courses could be provided before or during a surge in patient attendances to build resilience in HCWs. Meditation courses have the potential to alleviate stress in hospital staff and are an economical option. Cognitive Behavioural therapy may also be an option for selected healthcare workers, with options to adopt to virtual sessions. Sole debriefing sessions are not recommended. The need for evidence based psychological intervention research remains greater than ever.

## Data Availability

All data produced in the present study are available upon reasonable request to the authors

## Appendix

**Supplementary Table 1:**
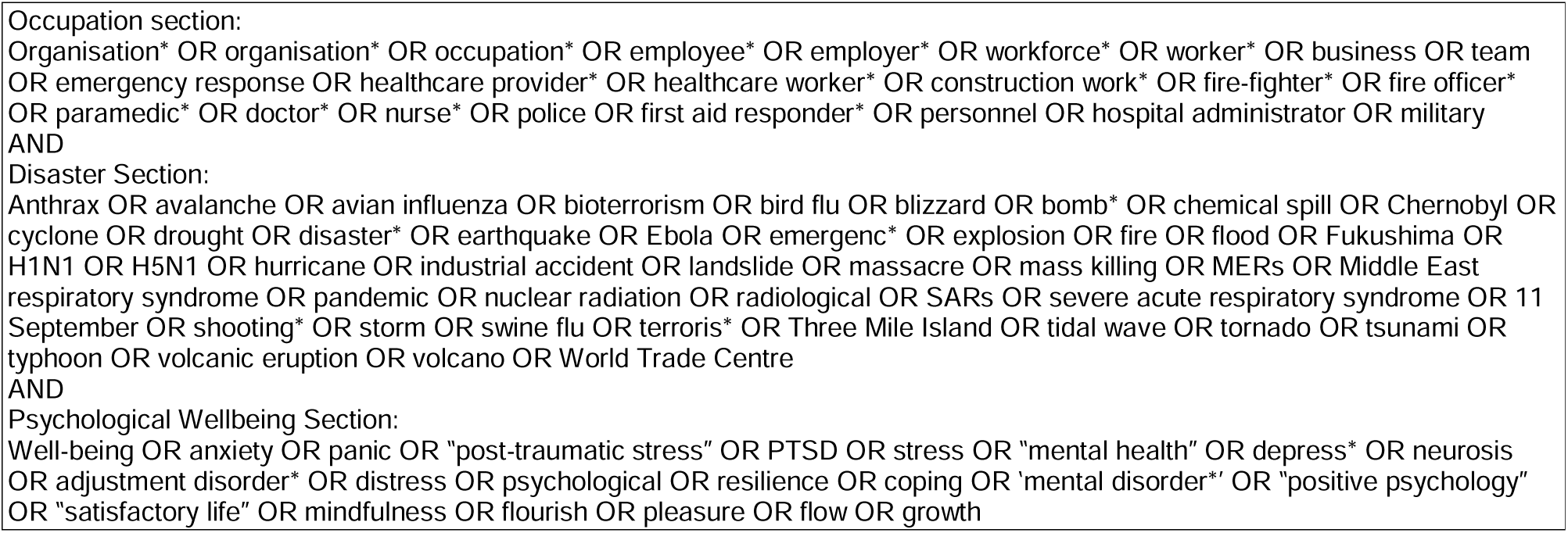
Search Strategy.

## References

1. Kisely, S., Warren, N., McMahon, L., Dalais, C., Henry, I. & Siskind, D. 2020. Occurrence, prevention, and management of the psychological effects of emerging virus outbreaks on healthcare workers: rapid review and meta-analysis. BMJ, 369, m1642.

2. Choudhury, T., Debski, M., Wiper, A., Abdelrahman, A., Wild, S., Chalil, S., More, R., Goode, G., Patel, B. & Abdelaziz, H. K. 2020. Covid-19 Pandemic: Looking After the Mental Health of our healthcare workers. J Occup Environ Med. 2020 Jul;62(7):e373–e376.

3. Muller, A. E., Hafstad, E. V., Himmels, J. P. W., Smedslund, G., Flottorp, S., Stensland, S., Stroobants, S., Van De Velde, S. & Vist, G. E. 2020. The mental health impact of the covid-19 pandemic on healthcare workers, and interventions to help them: a rapid systematic review. Psychiatry Res. 2020 Nov;293:113441.

4. Gavin, B., Hayden, J., Adamis, D. & Mcnicholas, F. 2020. Caring for the Psychological Well-Being of Healthcare Professionals in the Covid-19 Pandemic Crisis. Ir Med J, 113, 51.

5. Schloemer, T. & Schroder-Back, P. 2018. Criteria for evaluating transferability of health interventions: a systematic review and thematic synthesis. Implement Sci, 13, 88.

6. Brooks, s. K., dunn, r., amlôt, r., greenberg, n. & rubin, G. J. 2018. Training and post-disaster interventions for the psychological impacts on disaster-exposed employees: a systematic review. J Ment Health, 1–25.

7. Iwakuma, M., Oshita, D., Yamamoto, A. & Urushibara-Miyachi, Y. 2017. Effects of Breathing-Based Meditation on Earthquake-Affected Health Professionals. Holist Nurs Pract, 31, 177–182.

8. Jones, N., Burdett, H., Green, K. & Greenberg, N. 2017. Trauma Risk Management (TRiM): Promoting Help Seeking for Mental Health Problems Among Combat-Exposed U.K. Military Personnel. Psychiatry, 80, 236–251.

9. Ke, Y. T., Chen, H. C., Lin, C. H., Kuo, W. F., Peng, A. C., Hsu, C. C., Huang, C. C. & Lin, H. J. 2017. Posttraumatic Psychiatric Disorders and Resilience in Healthcare Providers following a Disastrous Earthquake: An Interventional Study in Taiwan. Biomed Res Int, 2017, 2981624.

10. Aiello, A., Khayeri, M. Y., Raja, S., Peladeau, N., Romano, D., Leszcz, M., Maunder, R. G., Rose, M., Adam, M. A., Pain, C., Moore, A., Savage, D. & Schulman, R. B. 2011. Resilience training for hospital workers in anticipation of an influenza pandemic. J Contin Educ Health Prof, 31, 15–20.

11. Maunder, R. G., Lancee, W. J., Mae, R., Vincent, L., Peladeau, N., Beduz, M. A., Hunter, J. J. & Leszcz, M. 2010. Computer-assisted resilience training to prepare healthcare workers for pandemic influenza: a randomized trial of the optimal dose of training. BMC Health Serv Res, 10, 72.

12. Waelde, L. C., Uddo, M., Marquett, R., Ropelato, M., Freightman, S., Pardo, A. & Salazar, J. 2008. A pilot study of meditation for mental health workers following Hurricane Katrina. J Trauma Stress, 21, 497–500.

13. Reid, W. M., Ruzycki, S., Haney, M. L., Brown, L. M., Baggerly, J., Mescia, N. & Hyer, K. 2005. Disaster mental health training in Florida and the response to the 2004 hurricanes. J Public Health Manag Pract, Suppl, S57–62.

14. Difede, J., Malta, L. S., Best, S., Henn-Haase, C., Metzler, T., Bryant, R. & Marmar, C. 2007. A randomized controlled clinical treatment trial for World Trade Center attack-related PTSD in disaster workers. J Nerv Ment Dis, 195, 861–5.

15. Gershon, R. R., Gemson, D. H., Qureshi, K. & Mccollum, M. C. 2004. Terrorism preparedness training for occupational health professionals. J Occup Environ Med, 46, 1204–9.

16. Tehrani, N., Walpole, O., Berriman, J. & Reilly, J. 2001. A special courage: dealing with the Paddington rail crash. Occup Med (Lond), 51, 93–9.

17. North, C. S., Tivis, L., Mcmillen, J. C., Pfefferbaum, B., Cox, J., Spitznagel, E. L., Bunch, K., Schorr, J. & Smith, E. M. 2002. Coping, functioning, and adjustment of rescue workers after the Oklahoma City bombing. J Trauma Stress, 15, 171–5.

18. Wu, S., Zhu, X., Zhang, Y., Liang, J., Liu, X., Yang, Y., Yang, H. & Miao, D. 2012. A new psychological intervention: “512 Psychological Intervention Model” used for military rescuers in Wenchuan Earthquake in China. Soc Psychiatry Psychiatr Epidemiol, 47, 1111–9.

19. Kenardy, J. A., Webster, R. A., Lewin, T. J., Carr, V. J., Hazell, P. L. & Carter, G. L. 1996. Stress debriefing and patterns of recovery following a natural disaster. J Trauma Stress, 9, 37–49.

20. Boscarino, J. A., Adams, R. E. & Figley, C. R. 2005. A prospective cohort study of the effectiveness of employer-sponsored crisis interventions after a major disaster. Int J Emerg Ment Health, 7, 9–22.

21. Boscarino, J. A., Adams, R. E., Foa, E. B. & Landrigan, P. J. 2006. A propensity score analysis of brief worksite crisis interventions after the World Trade Center disaster: implications for intervention and research. Med Care, 44, 454–62.

22. Miller-Burke, J., Attridge, M. & Fass, P. M. 1999. Impact of traumatic events and organizational response. A study of bank robberies. J Occup Environ Med, 41, 73–83.

23. Seyle, D. C., Widyatmoko, C. S. & Silver, R. C. 2013. Coping with natural disasters in Yogyakarta, Indonesia: A study of elementary school teachers. School Psychology International, 34, 387–404.

24. Eid, J., Johnsen, B. H., Saus, E. R. & Risberg, J. 2004. Stress and coping in a week-long disabled submarine exercise. Aviat Space Environ Med, 75, 616–21.

25. Wald, J., Taylor, S., Asmundson, G. J., Jang, K. & Stapleton, J. A. Literature Review of Concepts: Psychological Resiliency. 2006.

26. Kunzler Am, Helmreich I, Chmitorz A, König J, Binder H, Wessa M, Lieb K. Psychological interventions to foster resilience in healthcare professionals. Cochrane Database of Systematic Reviews 2020, Issue 7. Art. No.: CD012527. DOI: 10.1002/14651858.CD012527.pub2. Accessed 25 May 2021

27. Heath, C., Sommerfield, A., & Von Ungern-Sternberg, B. S. (2020). Resilience strategies to manage psychological distress among healthcare workers during the COVID-19 pandemic: a narrative review. Anaesthesia, 75(10), 1364-1371.

28. Behan, C. (2020). The benefits of meditation and mindfulness practices during times of crisis such as COVID-19. Irish Journal of Psychological Medicine, 37(4), 256-258.

29. Rose, S. C., Bisson, J., Churchill, R. & Wessely, S. 2002. Psychological debriefing for preventing post traumatic stress disorder (PTSD). Cochrane Database of Systematic Reviews.

30. NICE 2018. Post Traumatic Stress Disorder: NICE Guideline [Online]. National Institute for Health Care Excellence. Available: https://www.nice.org.uk/guidance/ng116/chapter/Recommendations [Accessed 20/07/2020 2020].

